# Deep learning distinguishes focal epilepsy groups using connectomes: Feasibility and clinical implications

**DOI:** 10.1101/2023.02.09.23285681

**Authors:** Christina Maher, Zihao Tang, Arkiev D’Souza, Mariano Cabezas, Weidong Cai, Michael Barnett, Omid Kavehei, Chenyu Wang, Armin Nikpour

**Author notes:** These authors contributed equally (Chenyu Wang, Armin Nikpour).

## Abstract

The application of deep learning models to evaluate connectome data is gaining interest in epilepsy research. Deep learning may be a useful initial tool to partition connectome data into network subsets for further analysis. Few prior works have used deep learning to examine structural connectomes from patients with focal epilepsy. We evaluated whether a deep learning model applied to whole-brain connectomes could classify 28 participants with focal epilepsy from 20 controls and identify nodal importance for each group. Participants with epilepsy were further grouped based on whether they had focal seizures that evolved into bilateral tonic-clonic seizures (17 with, 11 without). The trained neural network classified patients from controls with an accuracy of 72.92%, while the seizure subtype groups achieved a classification accuracy of 67.86%. In the patient subgroups, the nodes and edges deemed important for accurate classification were also clinically relevant, indicating the model’s interpretability. The current work expands the evidence for the potential of deep learning to extract relevant markers from clinical datasets. Our findings offer a rationale for further research interrogating structural connectomes to obtain features that can be biomarkers and aid the diagnosis of seizure subtypes.

## 1. Introduction

Focal to bilateral tonic-clonic seizures (FBTCS) are a perilous form of seizures in focal epilepsy and are regarded as a primary factor in seizure-related injuries, severe cardiac arrhythmias, and sudden unexpected death [1, 2, 3, 4, 5]. The signature of FBTCS are seizures that begin in one brain hemisphere and spread rapidly without warning to the opposite hemisphere. The varying cortical and subcortical propagation patterns [6] are often dependent on the seizure onset zone [7]. The inherent limitations in prospectively identifying at-risk individuals are associated with the increased morbidity and mortality risk of FBTCS. Quantifying structural connectivity patterns may help distinguish patients likely to develop FBTCS from those who are not. A more precise diagnosis can inform personalised treatment pathways, disease prognosis, and improve patient care.

Diffusion-weighted MRI (dMRI) is an in-vivo neuroimaging technique that allows extraction of analytical features commonly used to examine the white matter in the brain [8]. White matter abnormalities observed in dMRI-derived metrics (such as fractional anisotropy and mean diffusivity) and graph theory measures have been linked to focal epilepsy and FBTCS [9, 10, 11, 12]. Structural connectomes derived from dMRI can be used to represent the presence and strength of connections between two discrete brain regions (represented as “nodes”) via the white matter tracts (represented as “edges”). The analysis of structural connectomes in focal epilepsy has yielded a range of network-based biomarkers that may aid pre-surgical selection and pharmacological treatment planning [6, 13, 14]. The combination of structural connectomes with functional data from electroencephalography (EEG) has revealed the influence of structure-function coupling in seizure dynamics [15, 16], seizure propagation [17], and postoperative seizure freedom [18]. Importantly, phenomenologic models have shown that structural alterations can give rise to FBTCS [19].

However, only one of the recent network-based analyses evaluated node abnormality in FBTCS [6]. Additionally, prior research predominantly focuses on generalised or temporal lobe epilepsy (TLE) populations [20, 21, 22, 23], prompting the need for further examination of FBTCS groups specifically. Moreover, limited clinical adoption of network-based connectivity biomarkers has been attributed to methodological inconsistencies and lack of clinician expertise in network analysis [24]. Further evaluation of the structural connectome could overcome such barriers by strengthening the evidence for clinically explainable connectivity biomarkers of specific seizure types.

Machine learning (ML) models may aid the adoption of network-based biomarkers in the clinical setting. Unsupervised machine learning algorithms have shown great potential for producing a data-driven classification of patients into subgroups and predicting surgical treatment outcomes [25, 26]. Deep learning models, considered a comprehensive iteration of machine learning [27], have recently been applied to structural connectome data to predict Parkinson’s disease [28], cognitive deficits [29] and epilepsy [30, 31], achieving promising outcomes. However, the relatively new approach of applying deep learning models to classify patients using their connectomes has not been applied in the context of FBTCS.

Additionally, though deep learning is a powerful ML technique, the search space provided by connectomes can be significant. Brain parcellation atlases contain anywhere from 84 regions in the Desikan-Killiany (DK) atlas [32] to 384 regions in the Atlas of Intrinsic Connectivity of Homotopic Areas (AICHA) [33]. Such a large number of regions results in over 3000 connections for the DK atlas and even more from other atlases. While fine parcellation atlases such as the AICHA have their benefits, they may also increase the inherent effect of noise per voxel in smaller regions (since one voxel in a smaller region is a greater percentage of that region than one voxel in a larger region), amplifying the potential noise effects in the results. Therefore, it may be advantageous for the deep learning model to partition connectomes by extracting a data-driven subset of nodes and edges that may be important to a given group.

Moreover, there is growing interest in multimodal image integration pipelines suitably designed for the clinical workflow [34]. The value of connectomics in unveiling network alterations in drug-resistant focal epilepsy was recently underscored [35]. Leveraging deep learning in combination with network neuroscience presents opportunities to identify salient features in high-dimensional imaging datasets [36]. A deep learning model which interrogates connectome data might improve diagnosis if seamlessly incorporated into an imaging analysis pipeline implemented directly in the clinical setting. However, the investment of resources in implementing such pipelines requires further initial evaluation of deep learning models to determine their ability to reveal clinically relevant information.

The aim of the current work was twofold. First, we sought to explore the feasibility of a deep learning model to identify, with reasonable accuracy, nodes and edges from the structural connectomes that were most important in classifying patients with and without FBTCS, and focal epilepsy from controls. Second, we explored whether the model could select nodes and edges that held clinically explainable characteristics of the patient subgroups that might guide further analysis or aid diagnosis. This proof of concept study may help quantify the value of structural connectome data in prospectively distinguishing individuals likely to have FBTCS from those who may not.

## 2. Methods

### 2.1. Participants and Data

Twenty-eight adults with focal epilepsy were recruited from the Comprehensive Epilepsy Centre at the Royal Prince Alfred Hospital (RPAH, Sydney, Australia). MRI was performed at the Brain and Mind Centre (Sydney, Australia). Inclusion criteria were adults diagnosed with focal epilepsy, aged 18-60, presenting without surgery, who were willing and able to comply with the study procedures for the duration of their participation. Exclusion criteria were pregnant women and individuals with intellectual disabilities. Twenty control participants, who had no prior neurological diagnosis, were also included in the study. Written informed consent was obtained from all participants before study participation. Ethical approval was obtained from the RPAH Local Health District (RPAH-LHD) ethics committee (RPAH-LHD approval ID: X14-0347). All research and methods were performed in accordance with the Declaration of Helsinki and the relevant guidelines and regulations prescribed by the RPAH-LHD ethics committee.

### 2.2. Image acquisition

Image acquisition was described previously [12]. Briefly, all scans were acquired on the same GE Discovery™ MR750 3T scanner (GE Medical Systems, Milwaukee, WI). The following sequences were acquired for each participant: Pre-contrast 3D high-resolution T1-weighted image (0.7mm isotropic) using fast spoiled gradient echo (SPGR) with magnetisation-prepared inversion recovery pulse (TE/TI/TR=2.8/450/7.1ms, flip angle=12); and axial diffusion-weighted imaging (2mm isotropic, TE/TR=85/8325ms) with a uniform gradient loading (*b*=1000s/mm^2^) in 64 directions and 2 *b* 0s. An additional *b*0 image with reversed phase-encoding was also acquired for distortion correction [37].

### 2.3. Image processing to obtain structural connectomes

The T1 images were processed and segmented according to the DK atlas [32], using a modified version of Freesurfer’s recon-all (v6.0) [38], alongside an inhouse skull-stripping tool (Sydney Neuroimaging Analysis Centre). The processed data for each participant was visually inspected by a senior neuroimaging analyst, and minor segmentation errors were manually corrected. A 5 tissue-type (5TT) image [39] was generated using MRtrix3 [40]. The T1 image was registered to the mean *b*0 image and the resulting transformation matrix was used to transform the 5TT image to the diffusion image. A schematic of the image processing pipeline is shown in Fig 1, a.

**Figure 1:**
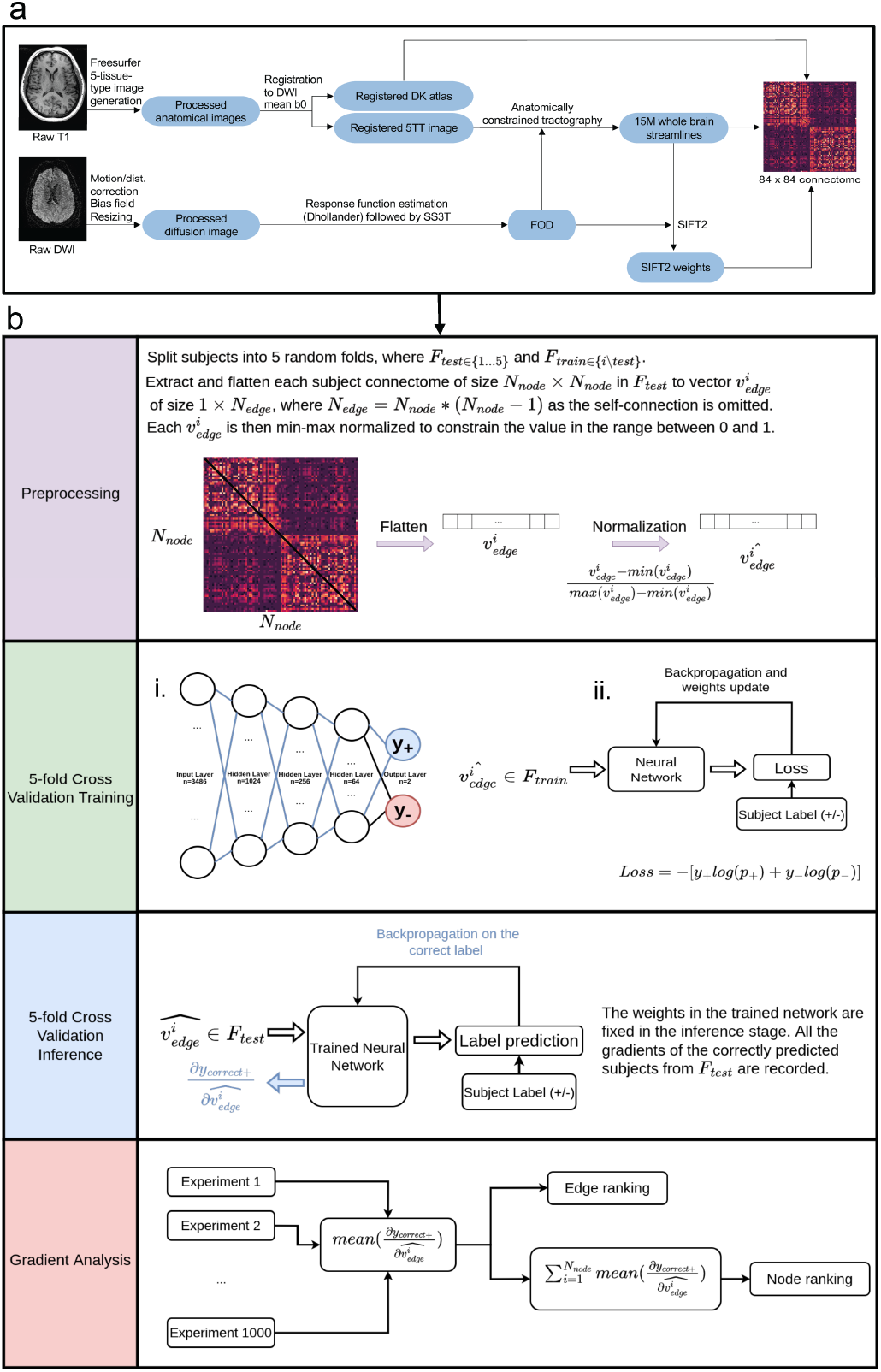
Image processing pipeline and deep learning model. In (a), the images were processed using our previously published pipeline[12], described in Sections 4.2 and 4.3. Next, the connectomes were fed into the deep learning model, shown in (b). The network architecture is shown in b,i. The network was trained with *F*_*train*_ using standard feed-forward and backpropagation (shown in b, ii). The network weights were then updated using the gradients calculated by cross-entropy loss and gradient analysis was conducted.

Diffusion image processing was conducted using MRtrix3 [40]. The diffusion pre-processing included motion and distortion correction [37, 41], bias correction using ANTs [42], and resizing to voxel size 1 mm isotropic. The *dhollander* algorithm [43] was used to estimate the response functions of the white matter, grey matter, and cerebral spinal fluid, from which constrained spherical deconvolution was used to estimate the fibre orientation distributions using MRtrix3Tissue (https://3Tissue.github.io), a fork of MRtrix3 [40]. The intensity of the white matter fibre orientation distributions was normalised [40], and used for anatomically constrained whole-brain tractography [44] (along with the registered 5TT image). The tractography protocols were as follows: 15 million tracks were generated, iFOD2 probabilistic fibre tracking [45], dynamic seeding [46], maximum length 300 mm, backtrack selected and crop at grey-matter-white-matter interface selected. For quantitative analysis, the corresponding weight for each streamline in the tractogram was derived using SIFT2 [46]. The streamlines and corresponding SIFT2 weights were used to create a weighted-undirected structural connectome using the registered DK parcellation image.

### 2.4. Experiment design

Classification experiments were conducted based on the participants’ classification labels. Participants were first classified into “All patients” and control groups. The “All patients” group was further split into those with (“FBTCS+”) and without (“FBTCS-”) FBTCS. Due to the relatively small sample size, two sets of experiments were separately designed for all participants (28 patients and 20 controls) and patient groups (17 FBTCS+ and 11 FBTCS-). For each set of experiments, the participants were randomly split into five folds for cross-validation to exploit the full dataset. Owing to the symmetric format of connectomes and to avoid data redundancy [47], only the upper triangle (shown in Fig. 1b) was used as input to the model, where each element represented the connection (“edge”) strength between two particular regions (“nodes”) in the DK atlas (total of 84 regions). The cross-validation was used to train and test 1000 experiments, each with randomised seeds for the initial network weights.

Finally, the edge values were normalised between 0 and 1 and flattened into a vector of 3486 values (with self-connections excluded).

### 2.5. Deep learning model

A neural network was chosen over traditional machine learning methods because it can learn more comprehensive non-linear feature representations from the input data. However, complex models with many parameters might overfit the data when given a small number of participants per group and render the interpretation meaningless. Therefore, to conduct an edge-wise and node-wise analysis of the results, a simple multi-layer perceptron (MLP) was selected due to its simplicity and suitability when using a flattened vector as input to the classification problem. A four-layer MLP was constructed with 1024, 256, 64, and 2 neurons (representing the two possible classes), respectively. The MLP was trained for 100 epochs with 1000 different seeds. The cross-entropy loss function optimised the network weights according to the participant labels. The network architecture is shown in Fig. 1, b, i.

#### Model analysis

The analysis was conducted only on the correctly predicted participants for each class of all 1000 5-fold cross-validation experiments to explore the region and connection importance in distinguishing the groups with different classification labels. The detailed analysis pipeline is shown in Fig. 1, b. Specifically, the back-propagation was applied to the correctly-predicted label for each participant with regard to their input values (i.e. the unique edges of the connectome). The corresponding gradients, representing the effect of a strong connection in the class prediction, were recorded and averaged for all the input node connections. Lastly, the corresponding connection weights for each node were summed to obtain the average gradient for each node (i.e. the strength of the node).

## 3. Results

### 3.1. Demographics

Twenty-eight patients (12 males, mean age and SD 40.32*±*12.38) were included in this study after meeting the inclusion criteria. Twenty age-matched healthy controls (5 males, mean age and SD 37.65*±*11.16) were also included. Participant characteristics are in Table 1.

**Table 1:**
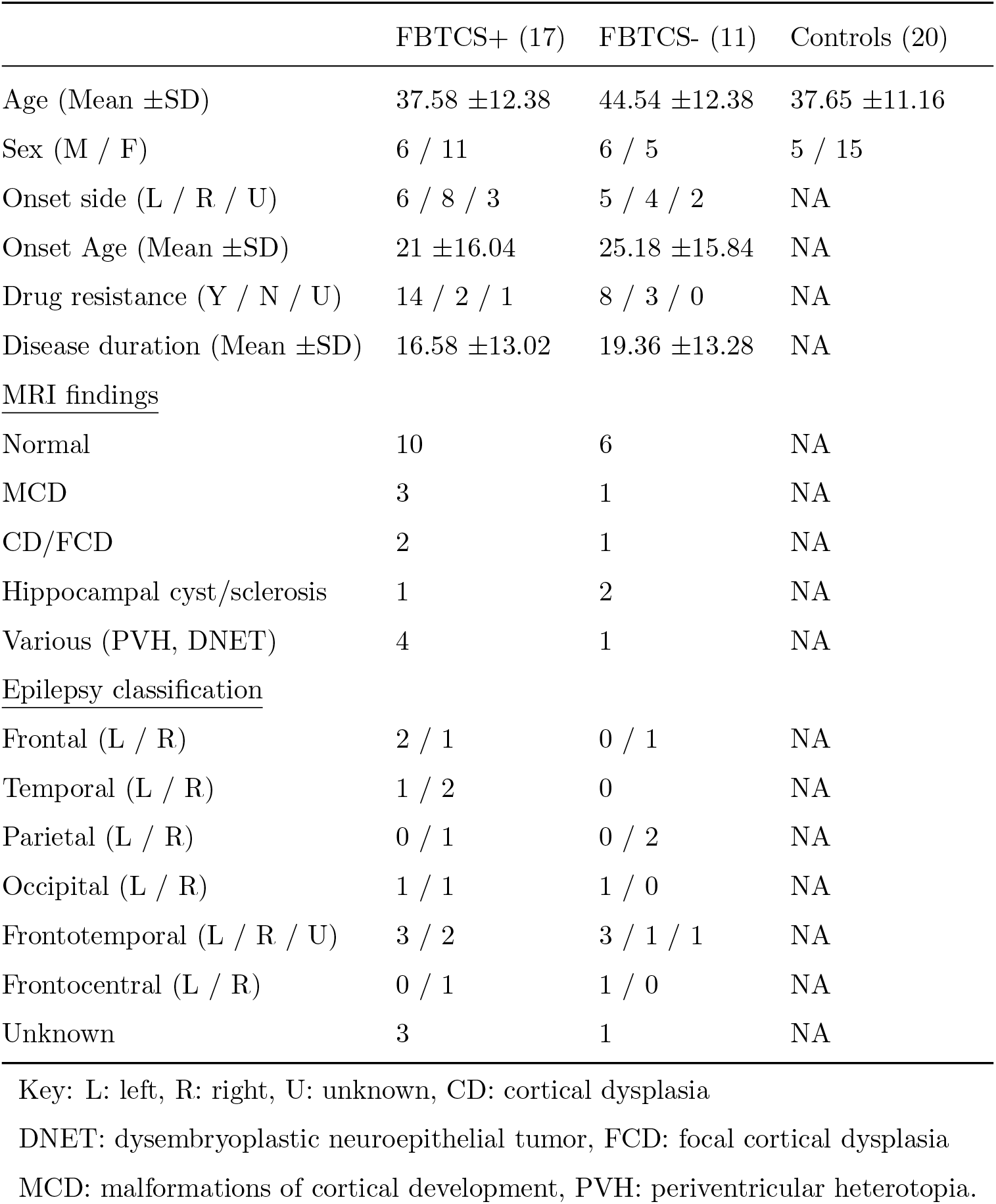
Participants’ demographic and clinical characteristics

### 3.2. All patients versus Controls

The best model from the trained network reached an accuracy of 72.92%. The model stratified important nodes and edges for each group by applying a gradient score which reflected the average overall probability of a node being important for classifying an individual to a given group. A positive gradient score indicated the positive contribution of a given node or edge to classifying a participant into a given group, with a higher score indicating more importance. Conversely, a negative gradient score indicated the negative contribution of a node or edge towards a participant’s classification into a group.

Notably, where a node or edge was assigned a positive gradient score for a given group, a negative gradient score was applied for that same node or edge for the opposite group, indicating the model’s ability to stratify each node or edge into only one of the two groups. The bar graphs in Fig 2 show the gradient score for the top 20 nodes and edges for the “All patients” group (edges: a, nodes: b). Here, the top 20 nodes and edges for the “All patients” group had a positive gradient score, yet the corresponding score for those same nodes or edges in the control group was negative. Although the model was first tested for classification accuracy between the “All patients” and control groups, the FBTCS subgroup classification results may bear more value for the field. Consequently, the following sections elaborate on the findings from the FBTCS groups’ analysis and the potential clinical implications.

**Figure 2:**
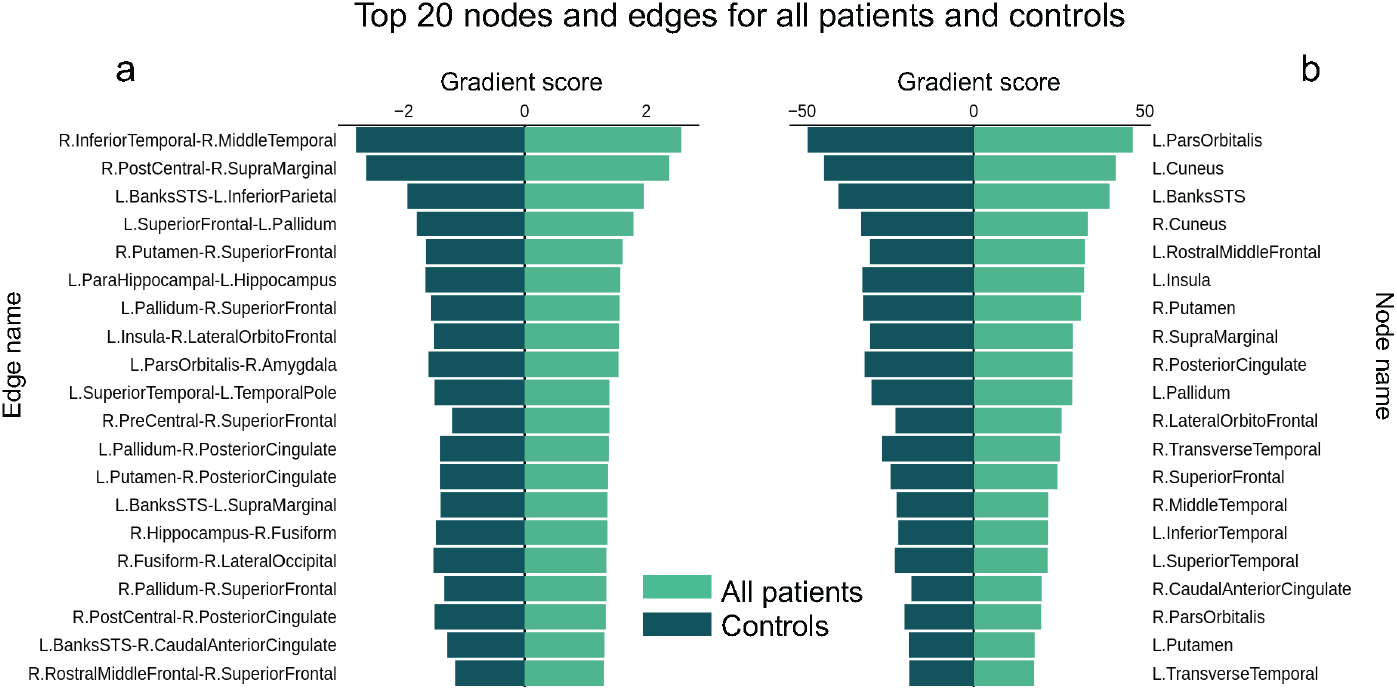
Gradient scores for the top 20 nodes and edges in the “All patients” group. The pyramid bar charts in (a) and (b) show the overall average gradient scores for the top 20 nodes and edges that demonstrated the greatest contribution to classification accuracy for the “All patients” group. Nodes were defined as per the DK atlas [32]. Where a node or edge was positive for the “All patient” group, the corresponding score was negative for the control group. Key: L: Left, R: Right.

### 3.3. Patient subgroups: FBTCS+ and FBTCS-

The best model trained and tested on the FBTCS+ and FBTCS-groups achieved a 67.86% accuracy score. To place the focus on the important nodes, only the top five nodes for each group are shown in Fig 3. Similar to the first classification test (“All patients” versus controls), a positive gradient score was assigned to nodes and edges consistently associated with accurate classification into a given group. Since FBTCS involves the propagation of a seizure from one hemisphere to another, the top edges (which imply strongly connected nodes) were of particular interest. The gradient scores for the top five edges for each group are illustrated in the bar charts in Fig. 4 (c and e). The top 10 edges for each group are illustrated in the brain images and chord diagrams in Fig 4, highlighting the key edge-based differences between the groups. Specifically, the model identified two edges (L.InferiorParietal-R.ParaCentral and L.IsthmusCingulate-R.Lingual) in the FBTCS+ group’s top 10 edges that crossed from one hemisphere to the other (illustrated in Fig. 4, a and b). In comparison, the FBTCS-group did not have any cross-hemisphere edges in the top 10 (illustrated in Fig. 4, d and f).

**Figure 3:**
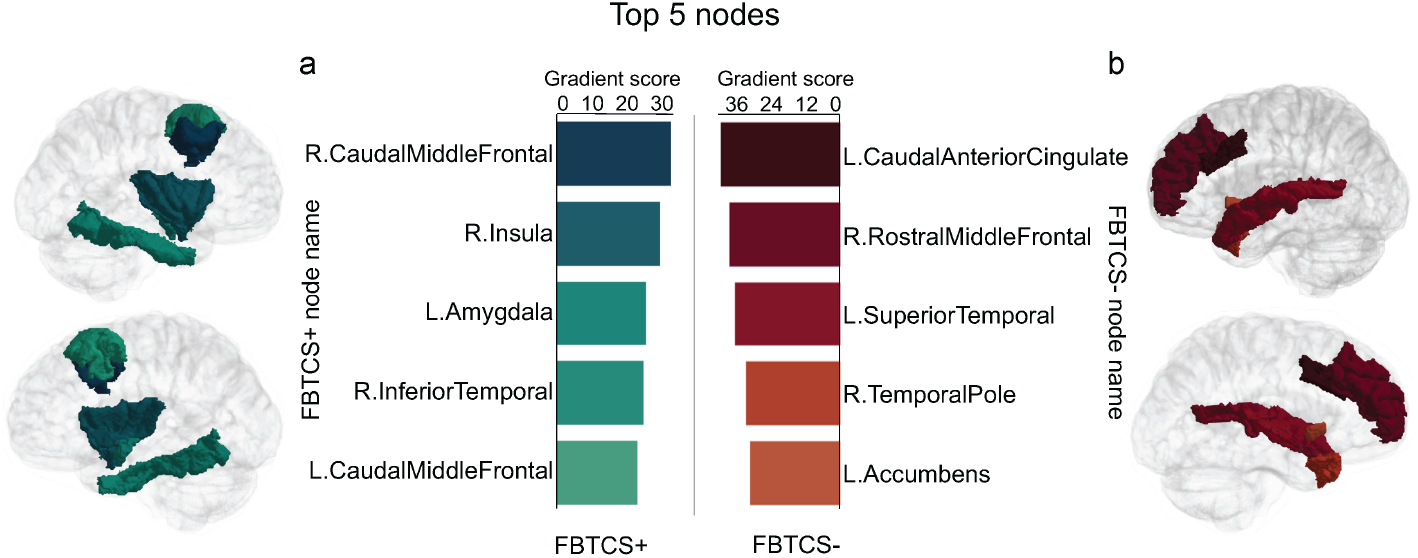
Top nodes for each FBTCS group. Nodes were defined as per the DK atlas [32]. The bar charts in (a) and (b) show the average gradient scores of the top five nodes that consistently demonstrated the greatest contribution to classification accuracy for each FBTCS group. Key: L: Left, R: Right.

**Figure 4:**
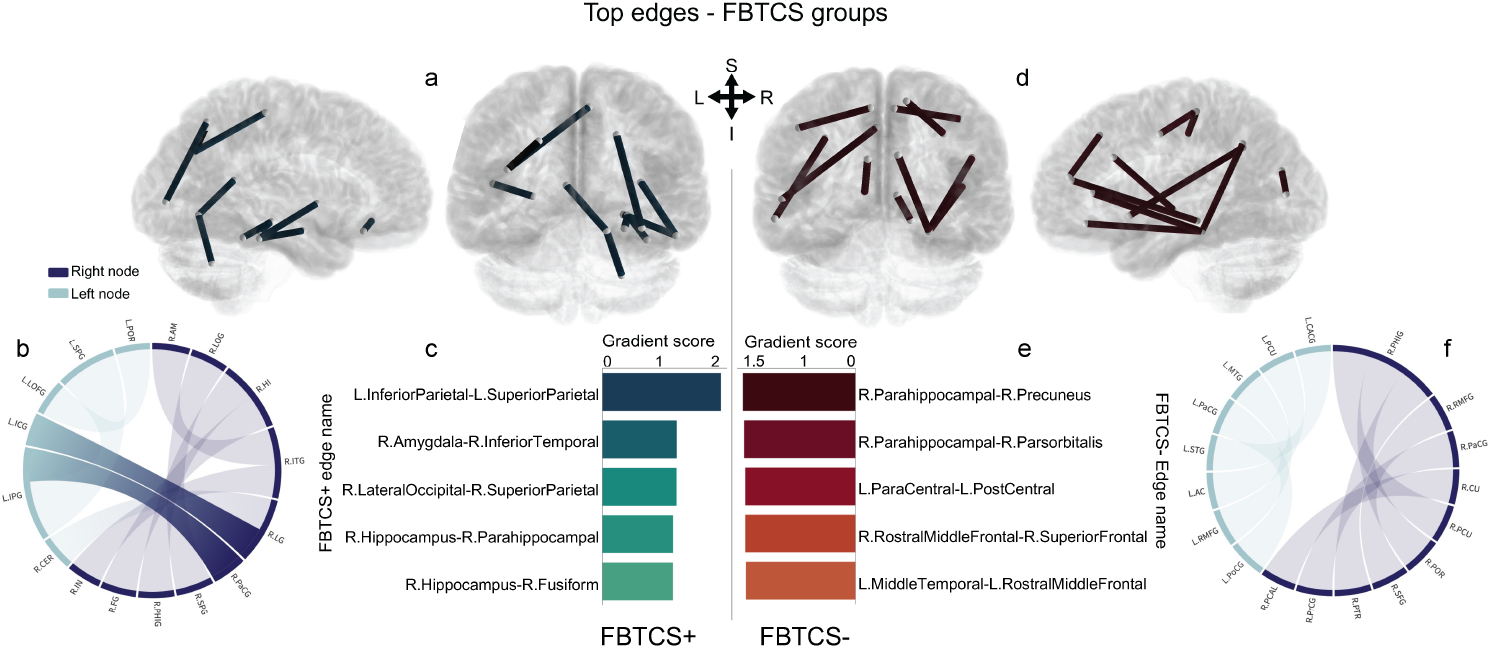
Top edges for each FBTCS group. The brain images and chord diagrams in (a and b) and (d and f) illustrate the main connectome differences between the FBTCS+ and FBTCS-groups. The bar charts in (c) and (e) show the top five edges for each group, which were also clinically relevant. Nodes were defined as per the DK atlas [32]. Specifically, the chord diagram in (b) shows that the FBTCS+ group had two cross-hemisphere edges its top 10 important edges, whereas the FBTCS-group did not. Key: L: Left, R: Right, AC: accumbens, AM: amygdala, CaCG: caudal anterior cingulate, CER: cerebellum, CU: cuneus, FG: fusiform, HI: hippocampus, ICG: isthmus cingulate, IN: insula, IPG: inferior parietal, ITG: inferior temporal, LG: lingual, LOG: lateral occipital, LOFG: lateral orbito frontal, MTG: middle temporal, PHIG: parahippocampal, PaCG: paracentral, PrCG: precentral, POR: pars orbitalis, PoCG: postcentral, PCAL: pericalcarine, PCU: precuneus, PTR: parstriangularis, RMFG: rostral middle frontal, SFG: superior frontal, SPG: superior parietal, STG: superior temporal.

To assess the model’s utility in identifying meaningful patient subgroup connections, the top 100 positive gradient scores for both grouping conditions were plotted according to nodal connections based in the left hemisphere (left to left), right hemisphere (right to right) and cross-hemisphere (left to right or right to left), as shown in Fig. 5. In the top 20 ranked edges for the “All patients” and control groups, the number of cross-hemisphere edges was relatively similar (shown Fig. 5, a, “Gradient rank - CR”). However, the top 20 ranked edges in the FBTCS+ group contained four cross-hemisphere edges (shown Fig. 5, b, “Gradient rank - CR”), compared to only one in the FBTCS-group.

**Figure 5:**
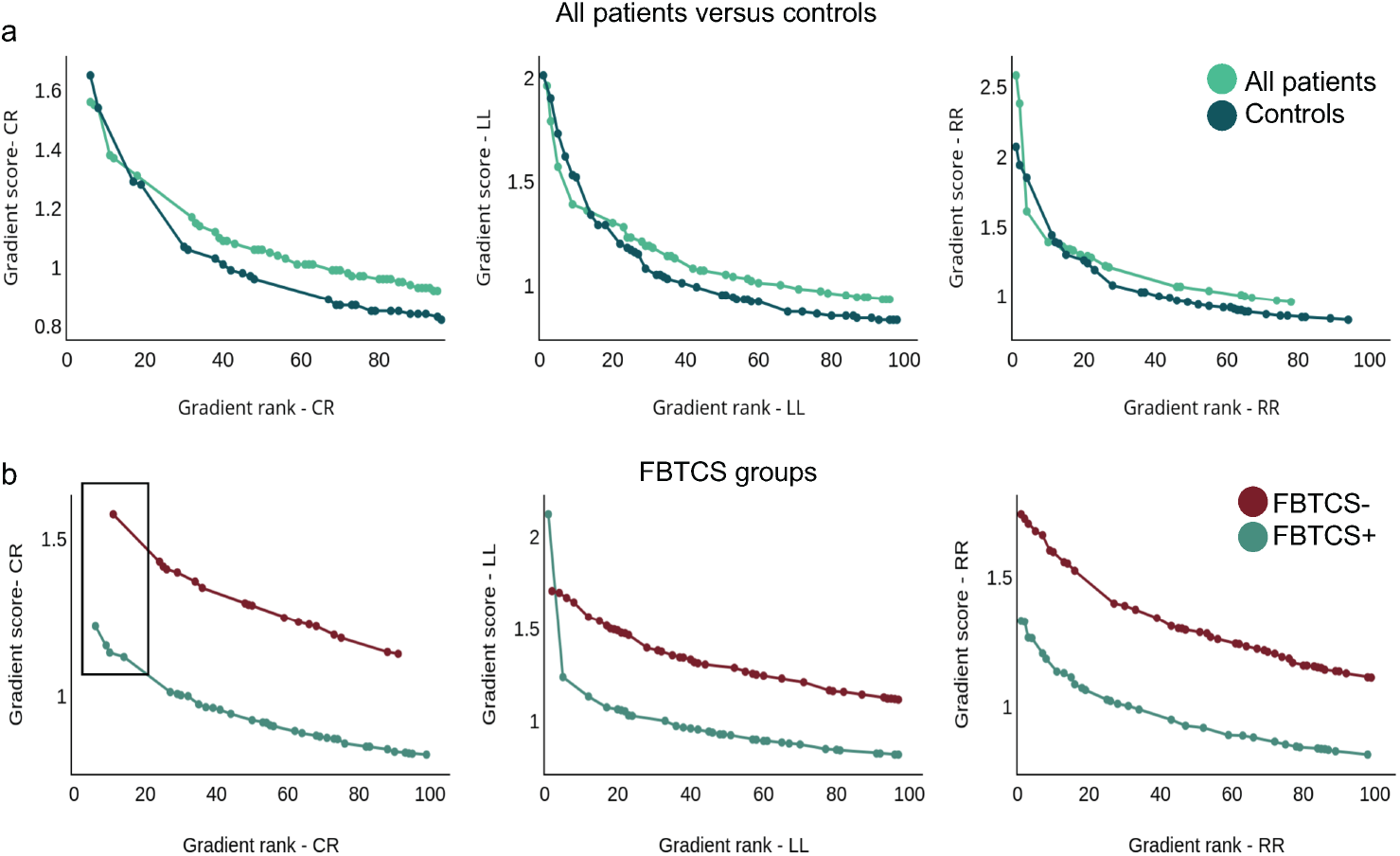
Top 100 edges for each group comparison. The top 100 ranked edges for each group were plotted as described in Methods Section 2.3. The “Gradient rank-CR” chart in (b) demonstrates the four cross-hemisphere edges in the FBTCS+ group compared to one cross-hemisphere edge in the FBTCS-group. Key: LL: left to left hemisphere, RR: right to right hemisphere, CR: cross-hemisphere.

## 4. Discussion

In this feasibility study, we applied a deep learning model to whole brain structural connectomes from controls and patients with focal epilepsy, who were further grouped by seizure type (FBTCS+ and FBTCS-). The model partitioned connectomes by assigning a gradient score to nodes and edges considered influential for classifying a participant into a given group. Results from the patient subgroups test identified connectome differences, disclosing unique nodes and edges relevant to each group’s clinical characteristics. This study provides initial evidence of the utility of our deep learning model as a tool to evaluate structural connectomes from patient subgroups. The clinical implications of the results are now discussed in the context of improving diagnosis with deep learning solutions.

First, our multi-layer perceptron model classified participants into their respective groups with similar accuracy to prior focal epilepsy research that applied different models and clinical questions. A recent work applied a supervised learning algorithm to classify the whole brain structural connectomes of controls and patients with TLE who were grouped according to seizure laterality, surgical outcome, and seizure freedom [31]. The DK atlas-based connectomes were combined with the fractional anisotropy (FA) diffusion metric, and MATLAB’s reconstruction independent components analysis algorithm [48] applied to reduce dimensionality [31]. The classification model achieved a receiver operating characteristic curve of 0.745 for the patients versus controls, 0.662 for left vs. right seizure onset, and between 0.77-0.80 post-surgical seizure freedom.

In a comparative work, Gleichgerrcht and colleagues [30] trained a deep network on whole brain structural connectomes and used 5-fold cross-validation to predict the postoperative seizure outcomes of 50 patients with mesial TLE. Unlike the current study, the authors introduced a binary ‘masking’ element to overcome the large number of parameters introduced by their chosen atlas (384-region AICHA atlas [33]). By feeding only the nodes above an arbitrary pre-specified into the deep learning model, the group achieved a classification accuracy with an 88*±*7 % positive predictive value for seizure freedom and amean negative predictive value of 79 *±* 8% for seizure refractoriness.

By applying the deep learning approach to a different classification problem in another epilepsy cohort, our findings expand the earlier evidence of the potential of deep learning solutions to probe connectomes and reveal clinically relevant information. Our “All patients” versus controls comparison resulted in a best model classification accuracy of 72.92% after 1000 repeated experiments. The subsequent training and testing of the model on our patient subgroups (FBTCS+ and FBTCS-) also yielded a better than random chance accuracy of 67.86% in the best model. The findings provide a proof of concept for our deep learning model as a preemptive tool to extract a subset of nodes and edges from large structural connectomes. In a clinical pipeline, the subset of nodes and edges could subsequently guide further connectome, tractography or graph metric analysis.

A second key result was our model’s disclosure of several nodes and edges known to be involved in focal epilepsy and FBTCS [9, 10]. The top five nodes and top 10 edges for each group were aligned to each group’s clinical characteristics in terms of the seizure onset zone and the structural and functional role of the region in seizure manifestation. Interestingly, since each group’s top nodes and edges were both ipsilateral and contralateral, they primarily represented the ratio of left versus right onset zones for each group.

Structurally, the top five nodes and edges in the FBTCS+ group contained primarily adjacent mesial structures, some anatomically closer to the lower subcortical regions. For instance, the fusiform gyrus borders the hippocampal gyrus and parahippocampal structures; the insula, amygdala, and inferior temporal regions are neighbouring regions, as are the parietal, occipital and cerebellum structures. The anatomical arrangement of structures in the FBTCS-group followed a similar proximity pattern. The frontal lobe houses the rostral middle and superior frontal structures adjacent to the pars orbitalis, while the temporal lobe houses the superior temporal and temporal pole nodes. The caudal anterior cingulate, precuneus, paracentral lobule, and postcentral gyrus are all neighbouring structures.

The contribution of the important nodes and edges to each group is buoyed by recent work exploring the influence of node abnormality in individuals with FBTCS. Sinha and colleagues reported that individuals with FBTCS+ had structural node abnormalities in subnetworks spatially correlated with their seizure onset zone [6]. The group showed that the node abnormalities in individuals without FBTCS tended to be localised to the temporal and frontal regions. In contrast, the significantly higher number of node abnormalities in the group with FBTCS group was more widespread and included the subcortical and parietal areas. A similar pattern was observed in our groups, as illustrated in Fig 3. Further, the five top-ranked nodes and edges in the FBTCS-group were consistent with prior findings of structural abnormalities in focal epilepsies without FBTCS [49, 10].

The functional perspective prompts several conceivable interpretations concerning the patient subgroups. First, prior works have linked several top nodes from the FBTCS+ group to secondarily generalised seizures using alternative imaging analysis techniques [50, 51]. In a study comparing 16 individuals with FBTCS to controls, voxel-based morphometry and resting-state functional MRI (rs-fMRI) were used to explore differences in structural and functional connectivity [50]. Compared to controls, the FBTCS group showed significantly increased functional connectivity from the left inferior temporal gyrus and left middle frontal gyrus to the thalamus. In contrast, decreased functional connectivity values were found between the thalamus and the right insula.

The mediating role of the thalamus in cortico-cortical communication in seizures that secondarily generalise has been analysed in rodent [52] and human [53, 11] studies. Reduced basal ganglia–thalamus network interaction is suggested to increase the propensity for secondary generalisation [53, 54]. These works suggest an impaired or reduced thalamo-cortical interaction, which may explain why the thalamus region did not receive a positive gradient score for classification into the FBTCS+ group. On the other hand, the thalamus received a positive gradient score for the FBTCS-group suggesting it may have an active role in inhibiting seizure propagation.

Second, the locale of the right pars orbitalis as a subregion of the inferior frontal gyrus offers the prospect of an electrophysiological inhibition mechanism in the FBTCS-group. The involvement of the right inferior frontal gyrus (rIFG) subregions in the basal ganglia and subthalamic nucleus response inhibition networks was evaluated in 31 participants using dMRI and rs-fMRI [55]. Reliable connections were identified between the pars orbitalis and the insula, putamen, caudate, and subthalamic nucleus, all regions involved in the stopping network. However, the current study design and data do not permit the unravelling of such mechanisms; this is an avenue for future research.

Third, the hippocampus and parahippocampal structures comprised one-half of the top 10 edges in both groups, a finding that may be explained by the groups’ clinical profile, as both had individuals with TLE. Hippocampal sclerosis is the most frequently observed pathology in drug-resistant TLE [56, 57]. Therefore, an alternative interpretation might consider the role of the hippocampus in individual variability of interhemispheric connectivity [58].

An operational hippocampal commissure has been shown in human epilepsy studies using depth EEG [59, 60]. Seminal work by Spencer and colleagues identified that in seizures arising in the hippocampus, contralateral neocortical involvement occurred with or after the contralateral hippocampus but never before [59]. In another study, the same group proposed the concept of a hippocampus that is initially functionally symmetric yet then directly or indirectly sustains an asymmetrical injury, i.e. one side is more damaged than the other. In recordings of the bidirectional interhippocampal seizure propagation time (ISHPT) from 50 individuals with TLE, a consistently longer ISHPT was emitted from the more affected hippocampus rather than to it [60]. The authors proffer that functional deterioration from neuronal cell loss can result in preferential propagation along the surviving efferent pathways unaffected by cell loss [60].

Returning to our patient groups, it is feasible that the degree of hippocampal aberration in the FBTCS-group creates a sufficient obstacle to neuronal recruitment and propagation of the abnormal electrical activity, such that it does not propagate contralaterally. Therefore, the engagement of the parahippocampal structure and its connection to the accumbens may be a preferential pathway in this group. In contrast, perhaps the relevance of the hippocampus to the FBTCS+ classification signals a level of cooperation between the hippocampus, amygdala, and cerebellum structures, which in turn supports a hypothesis of preferential commissural pathways in this group. In combination with prior seminal works, our findings support the theory of the structure-function relationship in FBTCS and highlight the value of a deep learning model in augmenting the clinical repertoire to improve diagnosis accuracy.

From a technical standpoint, our model may appeal to clinicians seeking turnkey solutions that utilise deep learning. The structural connectomes were generated using open-source software packages such as FreeSurfer [38] and MR-trix3 [40], and our deep learning model was developed in python code, removing the requirement for licensed software such as MATLAB. Therefore, one can envisage the entire pipeline (connectome generation and model deployment) seamlessly implemented in a hospital ward, enabling the prospective assessment and classification of new patients. Prospective evaluation of patients could inform clinical decision-making and feed into longitudinal studies.

Currently, diagnosing seizures such as FBTCS is a complex process requiring extensive observation and clinical and medical information. A clinician relies on patient and eyewitness descriptions if such seizures do not occur during the initial standard ward observation period. However, patient-reported histories have yielded inconsistent seizure classification at a rate of approximately 25% [61] due to onset signs being missed by the patient’s carer or the patient’s capacity to describe their seizure [62]. The benefits of a prospective classification into the FBTCS group include warning clinicians and carers to monitor precisely for such seizures, the potential to reduce instances of delayed diagnosis or misdiagnosis, and allowing tailored treatment that considers the possibility of such seizures. As evidenced by prior work, classification results could also inform patients’ presurgical candidature, including the likelihood of achieving postoperative seizure freedom. In that sense, fitting deep learning models into automated software that can be a precursor to further clinical investigation would enable an efficient and comprehensive diagnosis journey for the patient. With additional verification, the highest-scoring nodes could potentially act as a predictive biomarker, indicating regions requiring greater examination.

As with any deep learning approach, our results demonstrate the optimal level of accuracy attainable from the dataset and network architecture and must be interpreted as such. The modest sample size of some groups introduces the potential for overfitting and may restrict the generalisability of the findings, a concern shared by previous works [30, 31]. Additionally, the study and model design did not account for clinical variables like onset, node or edge hemisphere, which may limit the extent of interpretation. However, increasing the variables in a modest sample size may not be statistically valid. Further, our results must be considered in light of the DK atlas parcellations, which govern the extrapolations made from the top-ranking connections. The exploration and comparison of different atlases were beyond the scope of the current work, yet this is an important direction for future research. Our data collection is ongoing, with the view to address the dataset-related limitations and further validate the model. Future research will improve the model’s generalisability and accuracy by prospectively classifying new patients and comparing that classification with follow-up data in a longitudinal study design.

In conclusion, we applied a deep learning model to structural connectomes to classify patients from controls and individuals with and without FBTCS. The model identified important nodes and edges for classifying patients into a given group with a promising degree of accuracy given the modest sample size. The results from the patient subgroup classification were explainable from both the structural and functional perspectives, suggesting such a model would be valuable in improving the prospective diagnosis of FBTCS. The subsets of nodes and edges could also guide further patient-specific analysis. This work emphasises the potential of deep learning models designed to be clinically implemented as a tool to aid patient diagnosis. There is an opportunity to conduct a prospective classification of new patients, with a follow-up to determine whether the prediction was correct. Future research could focus on improving the model accuracy through training from a larger dataset and possibly including additional clinical variables.

## Data Availability

The datasets generated during and/or analysed during the current study are not publicly available because they are from RPAH patients, and access is only authorised for individuals named on the approved ethics application.

## 5. Acknowledgements

The authors acknowledge all staff at the Comprehensive Epilepsy Centre at the RPAH, particularly Mrs Maricar Senturias (RN/ACNC Epilepsy), who assisted with patient recruitment. The authors acknowledge the radiology staff at i-MED Radiology for their assistance with obtaining the MRI data.

## 8. Data availability

The datasets generated during and/or analysed during the current study are not publicly available because they are RPAH patients, and access is currently only for authorised individuals named on the approved ethics application. However, de-identified data can be made available from the corresponding author on reasonable request, subject to approval from the relevant governing ethics entities at the RPAH and The University of Sydney.

## 9. Code availability

The code for the deep learning model can be made available upon request.

## 10. Author statement

CM, CW, AN: Conceptualisation; CM, ZT, CW, AN: Methodology; CM, ZT, MC: Formal analysis; CM, ZT, CW, AN: Validation; CM, ZT, AD, MC: Software; CM, ZT, AD: Data curation; CM, MB, CW, AN: Resources; CM, ZT: Visualisation and Writing - original draft; WC, OK, CW, AN: Supervision; All authors: Writing - review/editing.

